# Dissociation of motor control from motor awareness in awake sleepwalkers: An EEG study in virtual reality

**DOI:** 10.1101/2020.11.17.20233072

**Authors:** Yannick Rothacher, Anh Nguyen, Evdokia Efthymiou, Esther Werth, Christian R. Baumann, Bigna Lenggenhager, Peter Brugger, Andreas Kunz, Lukas L. Imbach

## Abstract

Recent behavioral evidence indicates that awake sleepwalkers show dissociation of motor control and motor awareness in a virtual reality (VR) paradigm. Intriguingly, this dissociation resembles the nocturnal disintegration of motor awareness and movement during episodes of sleepwalking. Here, we set out to examine the neural underpinnings of altered motor agency in sleepwalkers by measuring EEG modulation during redirected walking in VR. Using this approach, we discovered distinct EEG patterns in awake sleepwalkers indicating facilitated dual tasking and salient habitual motor control as compared to healthy subjects. These observations provide electrophysiological evidence for the proposed brain-body dissociation in awake sleepwalkers. In conclusion, this study shows proof-of-principle that EEG biomarkers of movement in a VR setting might add to the diagnostic work-up of awake sleepwalkers.

## Short report

Patients suffering from parasomnia can enter a nocturnal dissociative state of body wakefulness and mind sleep [1]. Complex behaviors, including walking (‘sleepwalking’) and speaking, are executed in the absence of overt consciousness, while the brain remains in deep sleep [2]. The sense of agency - the awareness of initiating and executing motor action - is a key aspect of an individual’s bodily self [3–5].

In sleepwalkers, the mutual interaction between volitional control and bodily movements is temporarily decoupled during episodes of parasomnia. Sleepwalking can thus be conceptualized not only as an arousal disorder, but also as a *sleep disorder of motor agency*. In sleepwalkers, this dysfunctional brain state emerges exclusively during deep sleep, while awake sleepwalkers show normal performance in neurological or cognitive assessments. Therefore, diagnosis of parasomnia typically requires sleep examinations in specialized clinics. However, in a recent inspiring study, Kannape and co-workers [6] used a virtual reality (VR) paradigm to unmask altered motor awareness in sleepwalkers: Implementing a walking paradigm with deviated feedback, the authors investigated sleepwalkers while modulating motor control and perception. The core finding of the study was that sleepwalkers maintained more stable motor control and better motor awareness while walking under cognitive load, indicating a multi-tasking advantage. These findings imply that awake sleepwalkers might utilize different brain networks of motor control even during wakefulness.

We set out to test this hypothesis by measuring EEG activity in a VR paradigm analogous to the one used by Kannape et al. [6] to measure on-line electrophysiological biomarkers of motor control. To this end, we analyzed the central beta EEG power, a measure known to be reduced prior to and during actual and imagined motor behavior [7–10]. High beta power in precentral cortical areas has also been connected to lower cognitive load and more intrinsic (habitual) motor control [11]. In a similar vein, we recently found *high* beta activity in an invasive electrophysiological study during episodes of REM sleep parasomnia.[12] This fueled our hypothesis that beta power would be differentially modulated in awake sleepwalkers performing a motor task as compared to controls. Considering the superior motor performance under cognitive load for sleepwalkers in Kannape’s study, we expected higher beta activity (corresponding to more habitual motor control) in awake sleepwalkers, especially under dual task conditions (serial-7 subtraction task).

The experimental setup is depicted in Figure 1A, showing a participant in the virtual environment under variable feedback distortion that allowed for determining the subjective redirection threshold similar as in Kannape et al. In contrast to Kannape’s work, we implemented a first-person VR-perspective and a longer walking trajectory (Supplementary Table 1). Feedback distortion was achieved through the method of redirected walking [13]. Using this approach, the distinction between sub- or supra-threshold motor control can be interpreted as a measure of subjective motor awareness. This paradigm is an extension of the classical paradigm for measuring hand agency by Fournet and Jeannerod [14]. To increase the generalizability of our approach, we also determined redirection thresholds in an upper limb task (for details on the experimental procedures see Appendix).

**Figure 1:**
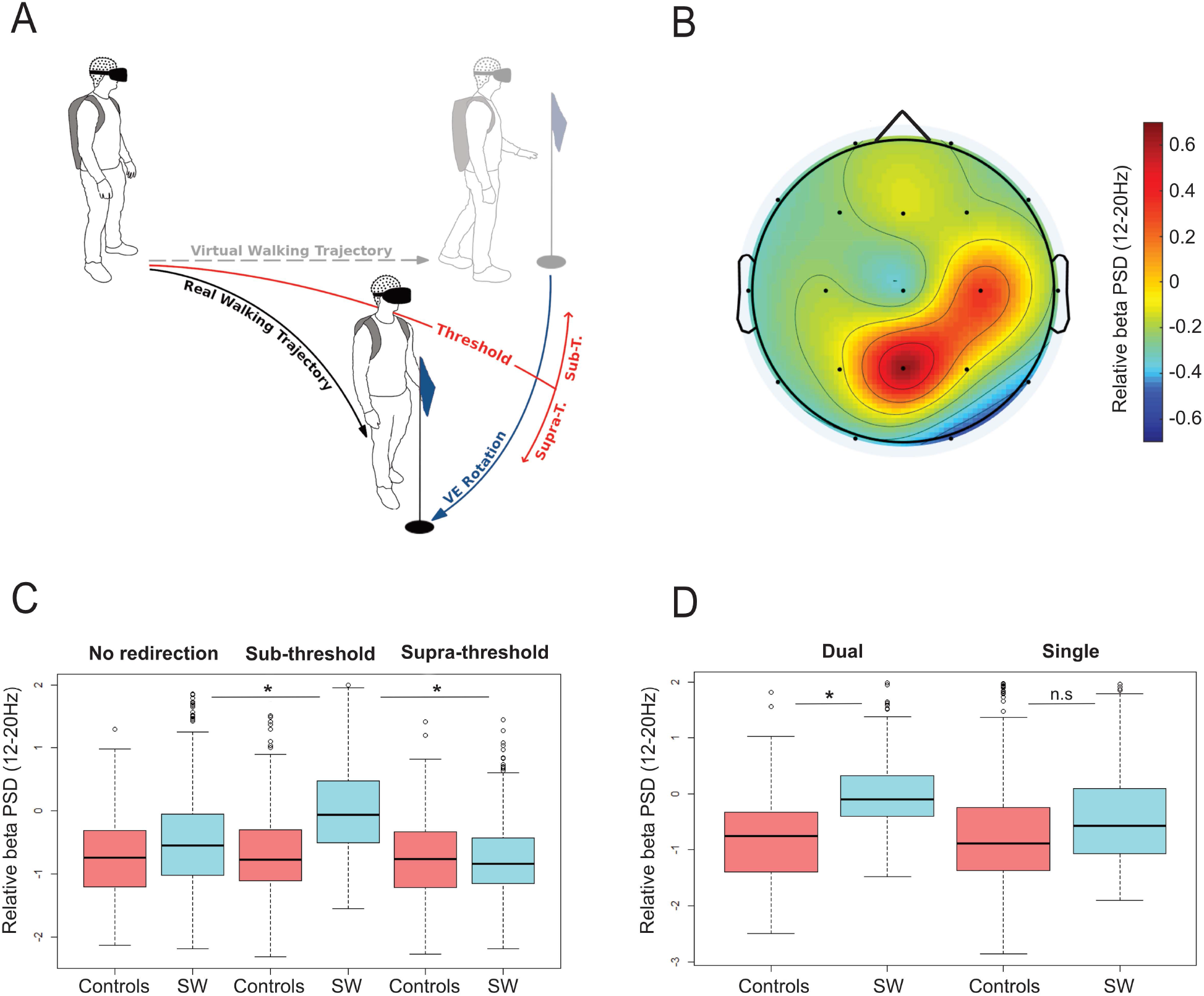
Experimental setup and main results. (A) Illustration of a walking trajectory manipulation through redirected walking with simultaneous measurement of surface EEG. Alternative forced choice was used to determine the individual redirection threshold. For trajectories below the threshold, participants are unaware of redirection (sub-threshold), while they are aware of redirection above (supra-threshold). (B) Mapping of low beta EEG power (13-20Hz) during redirected walking in sleepwalkers (relative to control subjects). Positive values indicate higher beta power for sleepwalkers (significant differences in C3, C4 and Pz). Electrode positions according to the 10/20 system are shown as black dots (with linear interpolation on a Cartesian grid). (C) Beta power relative to baseline (resting condition) during redirected walking is higher for sleepwalkers (SW) in the sub-threshold condition (middle panel, significant interaction between group and condition p<0.005). Post-hoc comparison showed significant differences for sleepwalkers in sub-threshold as compared to both other conditions (* p<0.005), but not for controls (n.s.). (D) Beta power relative to baseline (resting condition) is higher in sleepwalkers under dual task condition (significant interaction, * p<0.05) Post-hoc comparison shows a significant difference between sleepwalkers and controls only in dual task condition (* p<0.05). Beta PSD was normalized to pre-experiemental resting condition (set to zero) in C and D.

Our analysis focused on (i) reproducing the findings of Kannape et al. on a behavioral level and (ii) investigating the modulation of central beta power during the experimental procedures.

As in the preceding study [6], walking accuracy, velocity and subjective redirection threshold was not significantly different between the two groups (supplementary Table 1). However, in contrast to Kannape et al., we found no significant interaction between the two groups and conditions (cognitive load), neither in redirection threshold nor in motor performance (supplementary Table 1). On the other hand, the EEG analysis revealed significant beta power differences in sleepwalkers as compared to the control group (Figure 1B-C). First, the bivariate comparison between the groups showed overall higher beta power in sleepwalkers over the right central region (during re-direction to the left) in the lower beta band (Figure 1B). We focused therefore on the interaction of central beta activity with the experimental conditions (dual versus single tasks) and subjective awareness of redirection. Here, we found higher relative beta power in sleepwalkers under cognitive load (Figure 1D). Finally, we analyzed beta power in relation to the subjective redirection threshold during unbiased walking (no redirection), subthreshold redirected walking (unaware of redirection) and supra-threshold redirection (aware of redirection) to account for the modulation of motor control with individual motor agency. This analysis revealed significantly higher beta power in subthreshold redirected walking in sleepwalkers, but no difference in control subjects (Figure 1C). In other words, sleepwalkers shift towards more habitual motor performance (higher beta power) while being unaware of the redirection. Interestingly, we observed a similar pattern of altered motor control in subthershold condition also in an analogous upper-limb task (supplementary Figure 1).

While using a VR paradigm of redirected walking similar to Kannape’s study [6], we failed to reproduce their behavioral findings. This disagreement might be due to the first-person perspective implemented in our study, allowing for a more effective redirection in both groups. Alternatively, the use of a head-mounted display may have facilitated redirection over using a back-projection screen. In any case, the novelty we provide here is the electrophysiological evidence for the brain-body dissociation in awake sleepwalkers. In particular, we found that sleepwalkers shift efficiently to a more habitual motor performance by using intrinsic patterns of movement (under cognitive load and in subthreshold redirected walking, Figure 1C-D). This bias towards a more intrinsic motor control echoes the previous observation of better motor performance under cognitive load in Kannape’s work. Our findings complement these observations by documenting the sleepwalkers’ dissociation not only on a behavioral level, but also based on altered EEG biomarkers of motor control. As the same effects were also observed in an analogous task for the upper-limb (supplementary Figure 1), we argue that we revealed a characteristic higher-level property of altered motor control in sleepwalkers. The observed EEG changes in our study might thus provide insights in the neural underpinning for the altered motor control in sleepwalkers as observed by Kannape et al. In this way, both studies (albeit using different VR technologies) showed evidence for motor dissociation from awake awareness. Nevertheless, further studies are needed for a direct comparison of the different VR-paradigms on a behavioral and electrophysiological level.

In conclusion, our findings offer an intriguing electrophysiological explanation for the recurrent nocturnal episodes with unconscious motor action through habitual dual task control while simultaneously remaining in deep sleep. Finally, we provide further evidence that selective manipulation of motor control using a virtual environment can reveal significant differences in awake sleepwalkers as compared to control subjects, and might add to their diagnostic evaluation.

## Data Availability

Full access to patients/participants data are available upon reasonable request.

## Appendix 1 Methods and Procedures (online only)

### Participants

We recruited 15 patients (f: 7 and m: 8, mean age 30.6 y, range 22-47) with confirmed diagnosis of NREM parasomnia based on previous state-of-the-art sleep laboratory examinations (video-polysomnography). Additionally, all patients had a history of nocturnal episodes including sleepwalking. Eight patients had a positive family history for NREM parasomnia. Eight patients also reported injuries from nocturnal episodes. Based on the Edinburgh Handedness Inventory [1], 11 patients were right-handed, two patients were ambidextrous and 2 were left-handed. To avoid chronobiological effects on behavior and EEG, we randomly measured half of the patients (7 patients) in the morning hours (08.30) and the remaining half at noon (12:30).

For our control group, we included 15 age and gender matched healthy volunteers (f: 8 and m: 7) without sleep-related disorders. To match for handedness, we included two left-handed and 13 right-handed participants. The control group has a mean age of 26y (range 22-39, not significantly different as compared to the patients).

All participants signed an informed consent prior to starting the experiment. All experimental procedures were approved by the Cantonal Ethics Committee of Zurich (BASEC number: 2019-00195) and carried out in accordance with the ethical standards of the Declaration of Helsinki.

### Experimental procedure

In both groups, a non-invasive scalp EEG using the international 10/20 montage (23 surface electrodes) was applied. EEG activity was measured for 20 minutes at rest (prior to entering the VR environment) and during the entire experiment. We used a mobile EEG device to be able to measure EEG during motor tasks (Trex HD amplifier, Natus Neuroworks) with a standard EEG cap (GVB Multicap). EEG was sampled at 512 Hz. For the redirection threshold estimation, participants wore an Oculus DK2 HMD and were connected to an Intersense IS-1200 optical tracking system for 6 DOF head position tracking at 180 Hz [16] (Figure 1A). In redirected walking the virtual environment is rotated around the user, forcing him/her onto a curved pathway and consequently causing a mismatch between the visually perceived and the physically performed walking trajectory (Figure 1). Based on the individual redirection detection thresholds, one can distinguish sub- and supra-threshold motor control, which can be interpreted as a measure of subjective motor awareness.

For each participant, the redirection threshold was assessed under a control condition and a dual-task condition. In the control condition, participants started at one end of a 12 m × 6 m tracking area and found themselves in an empty virtual room with a red pillar 7.5 m in front of them. Redirection thresholds were determined in a two-alternative forced choice task (2AFC task). Participants were asked to walk straight to the red pillar for two consecutive trials. In only one of the two trials, a leftward redirection of a specific intensity was applied. To familiarize the participants with the virtual environment and the different paradigms, we performed six trial runs (3 with and without redirection) for both the single and the dual tasks condition. Training runs were balanced between groups to ensure the same level of habituation. Only one-sided redirection was applied to increase power and reliablity of the subsequent EEG analysis. After completion of the two trials, participants were asked in which of the two trials the redirection had taken place. Depending on the correctness of the answer, the tested redirection intensity was adapted in the next round. In total, each participant completed 50 rounds in each condition. The selection of the tested intensities and the final estimation of the detection threshold was done using the Bayesian-based adaptive threshold estimation procedure QUEST [17]. QUEST uses a psychometric function to model the probability of giving a correct answer for a specific redirection. The psychometric function starts at a guessing rate of 50% for low redirection gains and approaches a perfect detection rate for strong redirection gains. The detection threshold is classically determined as the stimulus intensity correctly detected in 75% of the cases. The individual detection threshold was determined for each participant and compared between groups. Furthermore, we used the individual redirection threshold for classification of each trial in subthreshold and supra-threshold walking. Trials with redirection without subjective awareness were labelled as subthreshold, whereas suprathreshold redirection refers to trial with subjective redirection awareness. This classification was then used to compare EEG biomarkers between walking trials with and without awareness of redirection (Figure 1A/C). In total, in each group 1450 trials were performed (725 trials for the control condition and 725 trials for the dual-task condition). Of the 1450 trials with redirection, 595 were in subthershold and 855 in supra-thershold condition with no significant difference between groups.

In the dual-task condition, the threshold estimation followed the same general procedure. However, participants were requested to perform a serial-7 subtraction task while walking towards the virtual target. Specifically, before starting each trial, participants were shown a randomly generated two-digit number on the screen. Starting with this number, participants had to continuously subtract the number seven (while walking) and report the solutions verbally. The starting values of the serial-7 subtraction task were set between 70-100 to make sure that no negative numbers were reached in a trial.

Each condition took approximately 30 minutes. The order of the two conditions was randomized and counterbalanced over participants. Before starting the redirection threshold estimation, participants performed a short series of training trials in both conditions in order to get used to the redirection procedure.

For the modulation of motor awareness and motor control for an upper limb task, we implemented a similar VR task as follows: Participants sat upright in front of a tablet (WACOM digitizer tablet PTH 651, 370×275 mm) wearing an HMD (Oculus DK2). In the HMD, participants were presented with a rectangular field, representing the drawing board from a bird’s eye perspective. Participants controlled a cursor on this field using a stylus on the digitizer tablet. For each trial, a starting and target position were indicated on the field. After moving the stylus to the starting position, the task for participants was to guide the pen straight to the target position on the other side of the rectangular field. Similar to the feedback distortion used in redirected walking, a redirection was induced into the movements of the cursor. Participants had to perform a curved drawing motion to counteract this manipulation and to guide the cursor straight to the target. The estimation of the detection thresholds of the induced feedback distortion was performed in the same fashion as for redirected walking (2AFC task). Because of the shorter amount of time needed for drawing compared to walking, more trials were performed. In total, 2320 trials were performed in each group (1160 trials for the control condition and 1160 trials for the dual-task condition). The dual task consisted of the same serial-7 subtraction task used in the redirected walking threshold estimation. To prevent participants from performing very quick drawing motions, a timer (visually represented as a clock) signalized participants the time in which they were supposed to reach the target position. Trials with a slower or faster drawing speed were labeled as invalid trials and were repeated.

### EEG post-processing

To quantify the modulation of spectral properties of the EEG signal during the motor task, we calculated the average power spectral density (PSD) during all experiments using a modified periodogram [18] approach (256 samples Hanning window, 50% window overlap, 512 Hz sampling rate). The full spectral analysis revealed a consistent beta peak in the low beta band (12-20Hz) over the central electrodes (C3/C4, Figure 1B). To adjust for inter-individual variability of the raw EEG amplitude and background beta power, we normalized the beta power during movement to the average beta power in the resting condition prior to movement initiation (baseline). This normalization also accounts for possible vigilance effects on the background beta levels across groups. Beta power was calculated in a common reference montage for mapping (Figure 1B) and in a bipolar montage from the region showing the most prominent modulation (C4-Cz) for the further multi-linear correlation analysis. No electrode clustering was performed for statistical analysis. We implemented a spectral-based artifact rejection (excluding epochs with gamma power (>30Hz) 3 times over the individual median. The number of rejected trials due to artifacts was not different between the groups (8% for sleepwalkers versus 9% for controls).

### Statistical analysis

We used R (R CoreTeam, 2012) to perform a linear mixed model analysis for the relationship between beta power (dependent variable) and group (sleepwalker/controls), task (dual/single) and redirection awareness (subthreshold/suprathreshold/control condition) as independent variables, respectively. Individuals were included as random variables. We controlled for age, gender and handedness and circadian effects (morning/afternoon measurement). All p-values were obtained by likelihood ratio tests. For post-hoc comparisons (data in Figure 1C-D), we applied the Satterthwaite method with Tukey adjustment. All analyses were repeated in the same way with the exclusion of the two left handed participants, showing the same significant effects as reported.

### Declaration of Interests

The authors declare no competing interests.

### Author contributions

Conceptualization, LLI, PB, AK, EW; Methodology, LLI PB AK BL.; Investigation, EE, AN, YR; Writing – Original Draft, LLI.; Writing –Review & Editing, all authors; Resources,CB, EW.; Supervision, LI, PB and AK.

**Supp.Figure 1.**
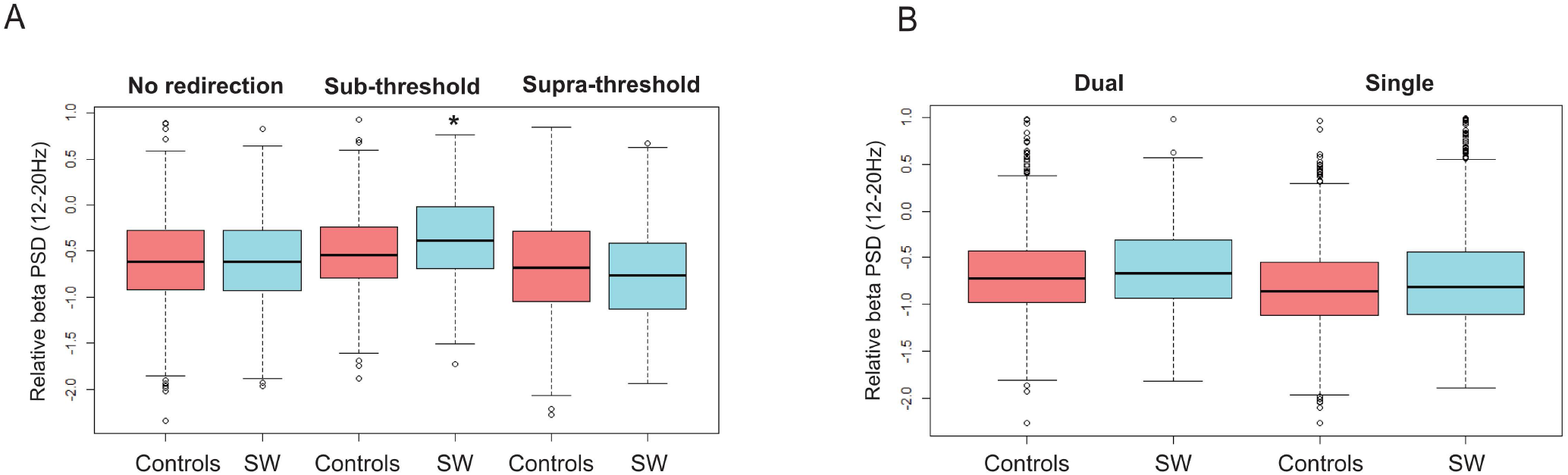
EEG beta modulation for the upper-limb task. (A) Beta power relative to baseline during redirected walking is higher for sleepwalkers (SW) in the sub-threshold condition (middle panel, significant interaction between group and condition *p<0.05). (B) Beta power relative to baseline is higher in sleepwalkers under dual task condition (significant interaction, p<0.05).

**Supplementary Table 1:**
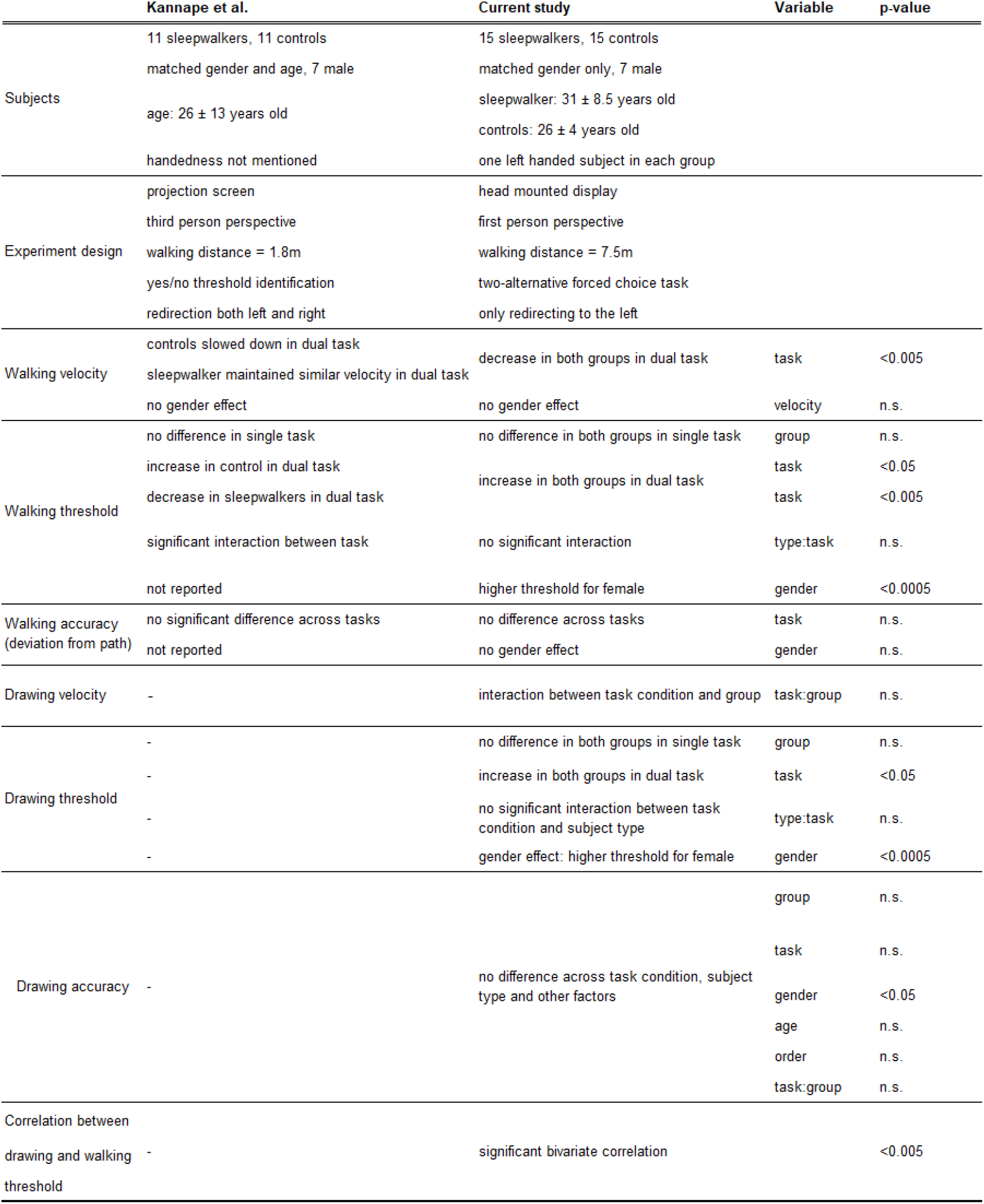
Comparison of behavioral effects in Kannape et al. [6] and the current study including velocity, redirection threshold, accuracy for the implemented walking and drawing paradigms. Analyzed variables are shown in column 4, tested interactions between variables are marked with a colon between variables (e.g. task:group). P values from a linear mixed model analysis for the dependent variable (column 1) are given in column 5.

## Acknowledgement

The study received no funding.

## Notes

### Competing Interest Statement

The authors have declared no competing interest.

### Funding Statement

none

### Author Declarations

Kantonale Ethikkommission Zurich

## References

1. Bassetti, C., Vella, S., Donati, F., Wielepp, P., and Weder, B. (2000). SPECT during sleepwalking. The Lancet 356, 484–485.

2. Arnulf, I. (2018). Sleepwalking. Curr. Biol. 28, R1288–R1289.

3. David, N., Newen, A., and Vogeley, K. (2008). The “sense of agency” and its underlying cognitive and neural mechanisms. Conscious Cogn 17, 523–534.

4. Castiello, U., Paulignan, Y., and Jeannerod, M. (1991). Temporal dissociation of motor responses and subjective awareness. A study in normal subjects. Brain 114 (Pt 6), 2639–2655.

5. Weiss, C., Tsakiris, M., Haggard, P., and Schütz-Bosbach, S. (2014). Agency in the sensorimotor system and its relation to explicit action awareness. Neuropsychologia 52, 82–92.

6. Kannape, O.A., Perrig, S., Rossetti, A.O., and Blanke, O. (2017). Distinct locomotor control and awareness in awake sleepwalkers. Curr. Biol. 27, R1102–R1104.

7. Imbach, L.L., Baumann-Vogel, H., Baumann, C.R., Sürücü, O., Hermsdörfer, J., and Sarnthein, J. (2015). Adaptive grip force is modulated by subthalamic beta activity in Parkinson’s disease patients. Neuroimage Clin 9, 450–457.

8. Schaller, F., Weiss, S., and Müller, H.M. (2017). EEG beta-power changes reflect motor involvement in abstract action language processing. Brain and Language 168, 95–105.

9. Kühn, A.A., Williams, D., Kupsch, A., Limousin, P., Hariz, M., Schneider, G.-H., Yarrow, K., and Brown, P. (2004). Event‐related beta desynchronization in human subthalamic nucleus correlates with motor performance. Brain 127, 735–746.

10. Klopp, J., Marinkovic, K., Clarke, J., Chauvel, P., Nenov, V., and Halgren, E. (2001). Timing and localization of movement-related spectral changes in the human peri-Rolandic cortex: intracranial recordings. Neuroimage 14, 391–405.

11. Bichsel, O., Gassert, R., Stieglitz, L., Uhl, M., Baumann-Vogel, H., Waldvogel, D., Baumann, C.R., and Imbach, L.L. (2018). Functionally separated networks for self-paced and externally-cued motor execution in Parkinson’s disease: Evidence from deep brain recordings in humans. Neuroimage 177, 20–29.

12. Hackius, M., Werth, E., Sürücü, O., Baumann, C.R., and Imbach, L.L. (2016). Electrophysiological Evidence for Alternative Motor Networks in REM Sleep Behavior Disorder. J. Neurosci. 36, 11795–11800.

13. Rothacher, Y., Nguyen, A., Lenggenhager, B., Kunz, A., and Brugger, P. (2018). Visual capture of gait during redirected walking. Sci Rep 8. Available at: https://www.ncbi.nlm.nih.gov/pmc/articles/PMC6299278/ [Accessed June 26, 2020].

14. Fourneret, P., and Jeannerod, M. (1998). Limited conscious monitoring of motor performance in normal subjects. Neuropsychologia 36, 1133–1140.

15. Oldfield, R.C. (1971). The assessment and analysis of handedness: the Edinburgh inventory. Neuropsychologia 9, 97–113.

16. Foxlin, E., and Naimark, L. (2003). VIS-Tracker: a wearable vision-inertial self-tracker. In IEEE Virtual Reality, 2003. Proceedings., pp. 199–206.

17. Watson, A.B., and Pelli, D.G. (1983). Quest: A Bayesian adaptive psychometric method. Perception & Psychophysics 33, 113–120.

18. Welch, P. (1967). The use of fast Fourier transform for the estimation of power spectra: A method based on time averaging over short, modified periodograms. IEEE Transactions on Audio and Electroacoustics 15, 70–73.

